# Value-for-money of school feeding programs in sub-Saharan Africa: A multi-country cross-sectoral analysis

**DOI:** 10.64898/2026.06.24.26356172

**Authors:** Francesco Ramponi, Tom Forzy, Isabelle Iversen, Sein Kim, Kathrin Durizzo, Pratibha Gautam, Solomon Tessema Memirie, Mizan Habtemichael, Fentabil Getnet, Kingsley Masamba, Willy-Marcel Ndayitwayeko, Mélance Ntunzwenimana, Boka Stéphane Kévin Assa, Bridget Rieth, Gloria Kamwi, Selma Ingula, David Uchezuba, Cesar Macuacua, Noimilto Mindo, Antonio Chioze, Emílio Tostão, Donald A.P. Bundy, Stéphane Verguet

## Abstract

**Introduction:** School feeding programs (SFPs) can contribute to address undernutrition in low- and middle-income country settings while can simultaneously improve education outcomes and deliver social and economic benefits. However, evidence on their comprehensive value for money (VfM) remains limited. This study models the multi-sectoral impacts of SFPs across education, health, social protection, and the local agricultural economy in five sub-Saharan African countries, providing benefit-cost ratio (BCR) and VfM estimates to inform policy decisions.

**Methods:** The analysis used secondary data from national reports, program budgets, and national household surveys. SFP impacts on education were assessed through changes in years of schooling and linked to lifetime income gains. Health impacts were quantified as averted anemia cases, converted into disability-adjusted life years (DALYs) and monetized using gross domestic product (GDP) per capita. Social protection benefits were measured as the in-kind value of school meals relative to household food expenditures, while local economic impacts were assessed through monetary transfers to smallholder farmers (SHFs) from local food procurement. BCRs were calculated considering education and health impacts, with additional VfM and expanded BCR estimates considering broader benefits and transfers.

**Results:** Across countries, SFPs were associated with a cumulative gain of 0.6 to 2.3 years of schooling per beneficiary. Moreover, reductions in anemia cases are expected to avert between 4 to 51 DALYs per 1,000 beneficiaries. The BCR for education and health ranged between 3 and 31. Meals provided covered up to 28% of annual household food expenditures for low-income families. Local food procurement generated economic transfers between USD 7 and USD 15 per beneficiary per year to SHFs.

**Conclusion:** SFPs demonstrate high VfM, offering significant educational, health, and economic benefits. Policymakers in sub-Saharan Africa should prioritize the expansion and scaling of SFPs to enhance educational attainment, improve health, and foster economic resilience, contributing to sustainable development.

## Introduction

Every day, more than 400 million school-aged children worldwide benefit from school meals through a variety of school feeding programs (SFPs), which can help prevent undernutrition and micronutrient deficiencies,^1^ thereby lowering the prevalence of nutrition-related diseases and reducing acute and chronic illnesses.^2^ The role of SFPs is particularly critical in low- and middle-income country (LMIC) settings, such as sub-Saharan Africa, where, on average, one in three children aged 5-14 suffers from nutritional deficiencies.^3^ In LMICs, nutrition activities in schools have proved to be effective, notably with observed reductions in anemia among children receiving school feeding for at least a year, especially among adolescent girls.^4–6^

Beyond health and nutrition, SFPs can also provide benefits toward the education, agriculture, and social protection sectors.^6,7^ Firstly, sick and undernourished children can miss school days, are more likely to struggle academically and exhibit higher dropout rates, poorer school performance, and higher levels of grade repetition.^8–12^ By improving the nutrition of children at schools, SFPs can improve school enrolment and attendance, and boost learning outcomes,^13–15^ with potentially significant educational impacts especially in impoverished areas, where families face significant food insecurity.^7^ Further, in the long term, enhanced academic performance can in turn positively impact on productivity as adults.^16,17^

Secondly, SFPs can benefit the local agricultural sector, particularly when feeding activities prioritize sourcing food from local farmers through home-grown school feeding (HGSF) programs, where the fresh food components of school meals are supplied through local purchases from smallholder farmers (SHFs).^18^ Through HGSF, SFPs can boost demand for locally produced food and create stable incomes for SHFs;^19,20^ by expanding predictable markets for agricultural produce, HGSF may also strengthen farmers’ incentives to adjust production decisions and increase the use of purchased agricultural inputs, such as fertilizer or pest-control inputs, where supply conditions allow.^21^ In turn, farmers may be incentivized to diversify crop production, contributing to a more resilient agricultural sector by reducing the risk of crop failure and improving the overall food supply.^22^

Thirdly, SFPs can act as a social safety net. Beneficiary households receive an in-kind transfer equivalent to the value of the food provided to the children attending school, which can significantly alleviate household food expenditures.^6^ The social protection component is particularly relevant in areas such as sub-Saharan Africa, where gross national income (GNI) per capita is about USD 1,650,^23^ and where an estimated 45% of the population lives below the poverty line.^24^ Meals provided through SFPs can therefore constitute a meaningful share of annual food expenditures, especially for low-income families, enhancing food security and household stability.^7^

Additionally, SFPs may serve as a platform to deliver additional health and education interventions.^1^ SFPs are increasingly becoming comprehensive multi-faceted interventions, with school-based health education interventions promoting healthy diet and hygiene practices, providing micronutrients supplementation, and administering deworming treatment.^25^ Such multi-component programs can generate further positive ripple effects; for example, when SFPs are coupled with water, sanitation, and hygiene (WASH) activities, additional benefits can occur, via reduced incidence of diarrheal diseases.^26^

Despite their demonstrated and potential benefits, the implementation of SFPs in LMICs often faces significant challenges, primarily due to insufficient funding, poor infrastructure, and organizational barriers.^1^ Comprehensive assessments of the costs and benefits of these programs would be essential to address such implementation challenges and provide governments with actionable insights on how to strengthen and potentially scale up school feeding activities. However, the evidence on the cost-effectiveness and value for money (VfM) of SFPs is limited,^27,28^ and research on their intersectoral costs and benefits remains scarce in LMICs.^29^

In this study, we address this gap by evaluating the VfM of SFPs across five sub-Saharan African countries: Burundi, Ethiopia, Malawi, Mozambique, and Namibia. We define VfM as a broader economic evaluation framework that considers program costs in relation to multiple categories of benefits, including monetized education and health gains, social protection effects, and impacts on the local agricultural economy. Within this framework, we calculate a range of benefit-cost ratio (BCR) estimates based on monetized education and health benefits, and complement these with additional indicators capturing household-level transfer value and local procurement effects. Together, these analyses provide policymakers with empirical evidence on the VfM of SFPs to support informed decision-making.

## Methods

SFPs across the five countries examined in this study vary in design, coverage, and implementation, and are managed by different entities, including national governments, international organizations such as the World Food Programme (WFP), and partnerships with non-governmental organizations (NGOs). The programs also vary in their operational approaches, with some employing centralized procurement while others prioritize HGSF models that source food from SHFs. To quantify program VfM, we first analyzed national reports, program budgets, and official documents provided by ministries of education, health, and agriculture, as well as WFP. Using these sources, we calculated costs per meal and costs per child per year for each program, considering frequency of meals, coverage, and the modality of implementation. Table 1 summarizes the key characteristics of the SFPs across the five countries; on average, the cost per meal was estimated at USD 0.18 (SD: USD 0.06). More information for each SFP is provided in webappendix A.

**Table 1.** Characteristics of school feeding programs, coverage and associated costs, in each country.

| Country | Implementation modality | Primary school children reached | Meals per year | Frequency of school meals and SFP duration | Cost per child per year (USD) | Cost per meal (USD) |
| --- | --- | --- | --- | --- | --- | --- |
| **Burundi** | WFP | 684,000 | 196 | Every school day, for 9 years | 51 | 0.26 |
| **Ethiopia** | Coexistence of various programs | 6,136,500 | 180 | Every school day, for 8 years | 38 | 0.21 |
| **Malawi** | Mary's Meals | 1,000,000 | 196 | Every school day, for 8 years | 8 | 0.04 |
|  | WFP HGSF | 600,000 |  |  | 20 | 0.10 |
|  | Nascent Solutions | 75,000 |  |  | 31 | 0.16 |
| **Mozambique** | PRONAE | 220,000 | 160 | Every school day, for 7 years | 32 | 0.20 |
|  | WFP HGSF | 80,000 | 163 |  | 36 | 0.22 |
| **Namibia** | NSFP | 457,000 | 190 | Every school day, for 7 years | 25 | 0.13 |
|  | WFP HGSF | 11,700 | 190 | Every school day (3 days HGSF, 2 days NSFP), for 7 years | 19 | 0.10 |

SFP: School Feeding Program; WFP: World Food Programme; HGSF: Home-Grown School Feeding; PRONAE: Programa Nacional de Alimentação Escolar (National School Feeding Program, Mozambique); NSFP: Namibian School Feeding Program. Every school day = 5 days per week, 9 months per year.

To estimate the educational impact of SFPs, we combined country-specific administrative and survey data on enrolment and educational attainment for SFP beneficiaries and non-beneficiaries.^30–36^ Across settings, impacts on enrolment, dropout, repetition, and promotion were used to derive changes in schooling trajectories and estimate the additional years of education attributable to SFP exposure. These estimates were computed using transition-based cohort simulations for Burundi, Mozambique and Namibia, and regression-based models for Ethiopia and Malawi.^37^ To estimate economic returns, we integrated data from national socioeconomic surveys and applied a standard Mincer equation, correlating additional schooling years with future income potential to calculate expected lifetime earnings increases.^38^

Based on literature estimates of SFP impact on reducing anemia,^4,5,39^ combined with country-specific regional prevalence data sourced from household surveys, we estimated the expected cases of anemia averted among children participating in SFPs across countries. We converted the averted anemia cases into disability-adjusted life years (DALYs) averted by accounting for both years of life lost (YLL) due to premature mortality, using estimates of increased mortality risk associated with anemia, and years lived with disability (YLD), calculated using standard disability weights for mild, moderate, and severe anemia.^40,41^ DALYs averted were converted into monetary values using the gross domestic product (GDP) per capita as a proxy for societal willingness-to-pay (WTP) to gain one year of healthy life.^42^ In a sensitivity analysis, we extended the health impact evaluation to include the potential prevention of stunting and diarrheal diseases (see details in webappendix B).

To quantify the social protection benefits of SFPs, we considered school meals as an indirect transfer to households. The monetary value of the transfer was estimated as the annual food cost of meals provided per beneficiary, assuming each beneficiary was associated with one household. This annual transfer value was compared with total annual household food expenditure using regional data from household surveys.^34–36,43–46^ Impacts were disaggregated by region and household expenditure quintile to account for variability in socioeconomic contexts.

We estimated the contributions of SFPs to the local agricultural economy in terms of the income generated for SHFs and farmer organizations through local food procurement. In each country, the proportion of food sourced locally was determined based on program-specific procurement data, when available, and the value of food purchased locally was calculated as the economic benefit directly accruing to SHFs and farmer organizations.

Finally, to assess the VfM of SFPs, we compared the program costs to the monetary benefits in the education and health sectors and calculated BCRs for education and health. Education benefits were valued in terms of increased lifetime earnings attributable to additional years of schooling, while health benefits were estimated as the monetary equivalent of DALYs averted. To present a more comprehensive VfM measure, we also incorporated additional benefits associated with social protection and local agricultural economy, resulting from the in-kind transfers to households and economic gains for SHFs. Costs are presented in 2026 USD, and we used a 3% annual discount rate to compute the present value of costs and benefits over the program duration.^47,48^

All computations were realized using R software (version 4.4.3). The study was determined non-human subjects research by the Institutional Review Board of the Harvard T.H. Chan School of Public Health (IRB23-0061).

## Results

Across the countries studied, participants in SFPs were associated with gaining an average of 1.3 (SD: 0.7) additional years of schooling compared to non-beneficiaries. The gains in schooling ranged from 0.6 to 2.3 years across countries, depending on the specific settings and features of the programs. Detailed country-level results are reported in webappendix C and elsewhere.^37^ Regarding health benefits, SFP participants were expected to experience significant reductions in anemia cases compared to non-beneficiaries; moreover, additional beneficial effects on cases of diarrhea and stunting could be observed. When converted to DALYs averted, the reductions in anemia cases corresponded to between 4 (Malawi) and 51 (Burundi) DALYs averted per 1,000 beneficiaries over the full duration of program exposure. Including reductions in stunting and diarrhea cases, SFPs were associated with 35 (Ethiopia) and 89 (Burundi) DALYs averted (see webappendix C).

Comparing the total discounted cost per beneficiary over the full SFP exposure period to the monetary benefits in education (i.e., the net present value of incremental lifetime income gains) and health (i.e., the monetary value of DALYs averted over the same exposure period), the BCRs ranged between 3 (Burundi) and 31 (Namibia). In Table 2, the monetary values associated with the education and health impacts and the BCR estimates are illustrated for each country, indicating that for every USD 1 invested in SFPs, a minimum of USD 3 and a maximum of USD 31 are generated through improved educational and health outcomes, in Burundi and Namibia, respectively.

**Table 2.** Monetary value associated with educational and health benefits, and benefit-cost ratios.

| Country | Total discounted cost (USD) | Monetary benefits per beneficiary (USD) |  | Benefit-Cost Ratio (BCR) |
| --- | --- | --- | --- | --- |
|  |  | Education | Health |  |
| Burundi | 409 | 1,160 | 11 | 2.9 |
| Ethiopia | 273 | 2,172 | 6 | 8.0 |
| Malawi | 95 | 1,138 | 2 | 12.0 |
| Mozambique | 212 | 764 | 10 | 3.7 |
| Namibia | 158 | 4,811 | 31 | 30.7 |

Considering the in-kind transfer received by SFP beneficiaries, on average, the value of school meals provided corresponded to between 1% (minimum value, observed in Namibia) and 10% (maximum value, Burundi) of total annual food expenditures per household. For the lowest-income households (belonging to 1^st^ quintile of consumption expenditure) the transfer value amounted to a minimum of 2% of annual food expenditures (in Namibia) to a maximum of 28% (in Mozambique) (Figure 1). Regarding the effect on the local agricultural economy, local procurement of meals under the HGSF modality generated monetary transfers to SHFs ranging from USD 7 per beneficiary per year in Burundi, USD 8 in Ethiopia and Malawi, USD 11 in Mozambique, to USD 15 in Namibia. However, the extent of these benefits depended on the share of SFP activities implemented through HGSF, which accounted for approximately 40% and 27% of program activities in Malawi and Mozambique, respectively, and only 2% in Namibia.

**Figure 1.**
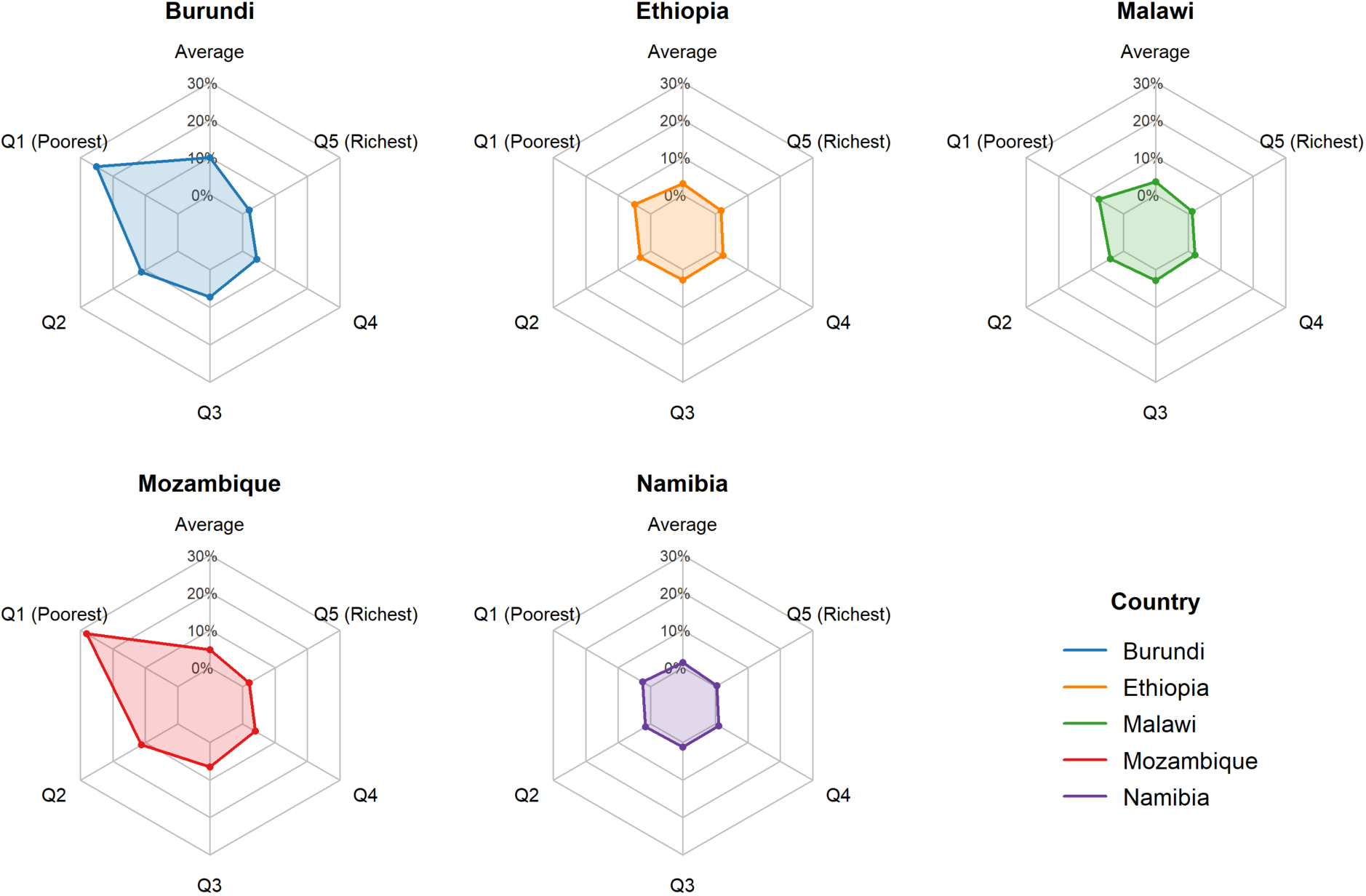
Value of the annual food transfer compared to annual household food expenditures, by consumption quintile.

The overall VfM estimates, calculated over the full SFP exposure period, are presented in Table 3. These include BCRs reflecting impacts on education and health, alongside additional effects on social protection, measured as the total discounted transfer to households per beneficiary, and on the local agricultural economy, quantified as transfers to local agriculture per beneficiary. Estimates for local agricultural effects reflect the current mix of HGSF and non-HGSF program modalities. We also report an expanded BCR, defined as the ratio of total monetized benefits across education, health, social protection, and the local agricultural economy to total program costs.

**Table 3.** Overall value-for-money of school feeding programs across sectors.

|  | **BCR**<br>**(education and**<br>**health)** | **Impact on**<br>**social protection**<br>**(USD)** | **Impact on**<br>**local agricultural**<br>**economy (USD)\*** | **Expanded BCR**<br>**(all benefits)** |
| --- | --- | --- | --- | --- |
| **Burundi** | 2.9 | 346 | 59 | 3.9 |
| **Ethiopia** | 8.0 | 145 | 59 | 8.7 |
| **Malawi** | 12.0 | 67 | 23 | 13.0 |
| **Mozambique** | 3.7 | 191 | 18 | 4.6 |
| **Namibia** | 30.7 | 140 | 2 | 31.6 |

\*Estimates reflect the current mix of home-grown school feeding (HGSF) and non-HGSF school feeding modalities.

## Discussion

Despite broad consensus on the critical role of SFPs in enhancing educational, health, social protection, and agricultural outcomes, particularly in LMICs in sub-Saharan Africa, evidence on their comprehensive value for money remains limited.^20,29,49^ In this study, we contributed to addressing this gap by quantifying the multi-sectoral impacts and overall value for money of SFPs across these dimensions. First, SFPs were shown to significantly improve children’s nutritional status, with estimated reductions in anemia among participants resulting in up to 51 DALYs averted. These estimated health impacts are likely conservative, as the analysis only included averted cases of anemia; when broader health effects such as reductions in stunting and diarrhea were considered, total health gains reached up to 89 DALYs averted (see webappendix C).^25^

Beyond health, addressing malnutrition through SFPs was also critical for education. The estimated increase of 0.6 to 2.3 additional years of schooling aligned with existing evidence and further underscored the importance of addressing food insecurity to support learning.^9,10,12,50^ Combining the monetized impacts on health and education, the BCR for SFPs ranged from 3 to 31, with consistently positive net monetary gains across all countries and higher returns observed in Malawi and Namibia.

Beyond their beneficial effects on health and education, SFPs also proved to be a critical social safety net, particularly for low-income families.^6^ The meals provided represented up to 25-30% of household food expenditures among the poorest quintiles in Mozambique and Burundi, reducing annual food costs and allowing households to redirect resources toward other essential needs. Of note, households with multiple children participating in the program may derive even greater social protection benefits compared to what estimated in our study. Finally, when relying on local food procurement, SFPs contributed significantly to local economic development.^22^ In Burundi and Ethiopia, transfers to SHFs and farmer organizations were estimated at approximately USD 59 per child per year. In Malawi, Mozambique, and Namibia, estimated transfers were USD 58, USD 67, and USD 96 per child per year, respectively; however, given the more limited implementation of HGSF in these settings, these gains were only partially realized at the national level.

A traditional cost-effectiveness analysis (CEA) focused solely on health maximization would have provided an incomplete assessment of the value of these programs. Estimated incremental cost-effectiveness ratios (ICERs) based on health impacts alone ranged from USD 1,662 (Malawi) to USD 54,661 (Ethiopia) per QALY gained across countries, with substantial variation depending on whether broader health effects such as reductions in stunting and diarrhea were included (see webappendix C). In contrast, the broader VfM framework applied in this study demonstrated high value for money, and captured substantial benefits particularly in the education sector, while also delivering significant health, social, and economic benefits. Even the more conservative VfM estimates in this study, which consider only education and health, indicated that SFP were associated with substantial BCRs.

In our analysis, we exercised caution in incorporating social protection and local agricultural effects into the main BCR estimates to avoid double counting, as these largely reflect resource reallocation rather than direct value creation. Nevertheless, they can be considered additional positive effects that enhance the overall value for money of SFPs from a redistributional perspective. Social protection benefits, in particular, may carry intrinsic societal value beyond their financial magnitude by alleviating poverty and improving household well-being. Similarly, transfers to SHFs should be interpreted as gross sales generated through SFP food procurement, rather than as net income gains, as the analysis did not account for production, transaction, or delivery costs borne by farmers or farmer organizations. These transfers may also be associated with broader spillovers not captured in this analysis, such as job creation and increased agricultural productivity. When social protection benefits and SHF procurement transfers were included alongside health and education benefits in expanded BCR estimates, the value for money of SFPs increased to up to 32.

Moreover, beyond their quantifiable impacts, SFPs may offer even greater value for money due to their broader contributions towards achieving the Sustainable Development Goals (SDGs). The health, nutrition, and education gains estimated in this study relate most directly to SDG 2 (Zero Hunger) and SDG 4 (Quality Education), by addressing food insecurity and malnutrition among school-aged children while supporting school participation and educational attainment. Beyond these two goals, HGSF programs can enhance local food markets and promote resilience, contributing to the development of sustainable food systems. SFPs may also serve as platforms for utilizing schools as hubs for delivering additional services, such as WASH and immunization programs. Additionally, school-based activities can foster social cohesion and community engagement, supporting peace and stability in fragile regions, such as Northern Mozambique.^51^

Capturing the full effects of SFPs was beyond the scope of the analysis, which focused on measurable impacts and adopted a partial equilibrium approach to estimate local economic effects due to limited data on agricultural productivity, production and transaction costs, and market dynamics. The analysis relied on secondary data sources, with important data gaps, particularly for health outcomes among school-aged children, and the absence of longitudinal evidence tracking long-term effects on education and health further limits the robustness of the estimates. Educational impacts were measured in terms of additional years of schooling, without accounting for changes in education quality. In addition, the analysis did not account for potential additional costs associated with increased educational attainment (e.g., further schooling), nor for cost savings resulting from improved health outcomes; these omitted components may partially offset each other. Finally, the combined effects of integrated interventions, such as SFPs delivered alongside WASH, deworming, or immunization programs, could not be assessed.

With this study, we provided new evidence on the multi-sectoral impacts and value for money of SFPs, demonstrating substantial benefits across education, health, social protection, and the local agricultural economy, alongside alignment with broader development objectives, including multiple SDGs. These findings highlight that single-sector approaches, such as traditional CEA focused on health outcomes, risk underestimating the full value of SFPs, whereas a broader benefit-cost analysis (BCA) and VfM framework provides a more comprehensive basis for policy decisions. Given these combined benefits, SFPs should remain a high priority for policymakers in sub-Saharan Africa. Continued expansion and scaling of these programs can enhance educational attainment, improve child health, and strengthen economic resilience, supporting sustainable development in the region.

Taken together, our findings highlight that approaches focused on a single sector, such as traditional CEAs centered on health outcomes, risk underestimating the full value of SFPs. In contrast, the broader BCA and VfM framework adopted in this study captures their multi-sectoral benefits and provides a more comprehensive basis for policy decisions. Given these multifaceted benefits, SFPs should remain a high priority for policymakers in sub-Saharan Africa, with continued expansion and scaling expected to improve educational attainment, child health, and economic resilience, contributing to sustainable development in the region.

## Data Availability

All data produced in the present study are available upon reasonable request to the authors

## Acknowledgements

We thank the Research Consortium for School Health & Nutrition for funding through an original grant by the Norwegian Agency for Development Cooperation (Norad), and follow-on support from the World Food Programme, USDA McGovern-Dole, the Rockefeller Foundation, the Bundesministerium für wirtschaftliche Zusammenarbeit und Entwicklung (BMZ), and the Novo Nordisk Foundation. The funders had no role in study design, data collection and analysis, decision to publish, or preparation of the manuscript.

Earlier versions of these findings were published as research working papers by the Research Consortium for School Health & Nutrition (https://www.lshtm.ac.uk/research/centres-projects-groups/research-consortium-for-school-health-and-nutrition).

