## Appendix for "Value-for-money of school feeding programs in sub-Saharan Africa: A multi-country cross-sectoral analysis"

### Webappendix A: School feeding programs in the sample countries

We provide below a brief overview of the various school feeding programs (SFPs) in the five countries selected for this analysis, covering: main activities; SFP coverage; meal frequency; source of food; and other activities included.

In Burundi, SFP activities are led by the World Food Programme (WFP), which provides locally sourced and imported nutritious meals, reaching approximately 40% of primary school children. The program integrates school meals with local agricultural support through school gardens and WASH training to improve hygiene practices and nutritional outcomes.^1^

In Ethiopia, SFPs are regionally tailored and provide fortified cereals, oil, and salt, served five days a week throughout the school year. Implemented by the Ministry of Education with support from WFP, the program reaches over 450,000 beneficiaries across seven regions. Fresh food pilots have also been introduced in select regions, integrating locally sourced vegetables and fruits into meals, along with complementary hygiene education.^2^

National SFP in Malawi is administered by the Ministry of Education in collaboration with the WFP and various partners, including for example Mary’s Meals and Nascent Solutions (considered in this analysis). The program provides supercereal (CSB+) and locally sourced foods, reaching approximately 30% of schools nationwide. It also supports local agriculture through the Smallholder Agriculture Market Support (SAMS) initiative, linking 34,000 farmers to the program. Additional activities include WASH training, education support, and agricultural development initiatives.^3^

In Mozambique, the National School Feeding Program (PRONAE) operates under a HGSF model, sourcing food from smallholder farmers and integrating additional interventions such as school gardens, deworming, and nutrition education. Managed by the Ministry of Education and supported by WFP, the program currently covers 340 schools, reaching over 220,000 children annually. PRONAE emphasizes local procurement and community involvement to bolster food security and promote agricultural sustainability.^4^

In Namibia, the National School Feeding Program (NSFP) is led by the Ministry of Education, Arts, and Culture, covering approximately 1,500 schools and approximately 500,000 students. The program provides centrally procured maize blend meals five days a week. A pilot HGSF model, involving 29 schools, integrates locally sourced diverse foods, benefiting smallholder farmers and enhancing local economic outcomes.^5^

### Webappendix B: Impact on health

**Figure B1**: Identification of studies considered in the health and nutrition impact analysis.

**Identification of studies via databases**

**Included**

**Screening**

**Identification**

Studies included in review:

n = 6

Records screened and assessed for eligibility:

n = 70

Records identified from:

n = 70

Records identified using search string:

‘school feeding’ OR ‘school meals’ OR ‘school meal’ OR ‘school nutrition’ OR ‘school lunch’

AND ‘anemia’ OR ‘anaemia’ OR ‘iron deficiency’

Records excluded: (n = 64)

Reason 1 (n = 25): Estimated the impact of SFP on other metrics

Reason 2 (n = 26): Did not estimate the impact of SFP

Reason 3 (n = 5): Latin America

Reason 4 (n = 2): Caribbean

Reason 5 (n = 2): Not in English

Reason 6 (n = 1): Europe

Reason 7 (n = 1): Targeting pre-schoolers

Reason 8 (n = 2): Asia

**Table B1**: Summary of the six extracted studies estimating the impact of school feeding on anemia and stunting.

| **Study** | **Year** | **Country** | **Type of Study** | **Intervention** | **Intervention details** | **Data collection year** | **Findings** |
| --- | --- | --- | --- | --- | --- | --- | --- |
| Neervoort et al. (2013)^6^ | 2013 | Kenya | Cross-sectional study | Feeding, education, and vitamin supplementation | 146 primary school children | 2009 | 55% reduction in anemia; 46% reduction in stunting |
| Abizari et al. (2014)^7^ | 2014 | Ghana | Cross-sectional study | School feeding program | 383 students aged 5-13 years | 2008 | No statistically significant effect on anemia or stunting |
| Adelman et al. (2019)^8^ | 2018 | Uganda | Cluster RCT | School feeding program and take-home ration | Adolescent girls aged 10-13 years | 2007 | 60% reduction in anemia |
| Walingo & Musamali (2008)^9^ | 2008 | Kenya | RCT | School lunch program | 320 students aged 10-12 years | 2008 | 45% reduction in severe stunting |
| Kwabla et al. (2018)^13^ | 2018 | Ghana | Cross-sectional study | School feeding program | 359 students aged 5-12 years | 2016 | No statistically significant effect on stunting |
| Hussein et al. (2023)^10^ | 2023 | Ethiopia | Comparative cross-sectional study | School feeding program | 936 primary school children | 2021 | 31% reduction in stunting |

RCT = randomized controlled trial.

*Stunting*

For the sensitivity analysis considering stunting, we completed a review of the literature and identified one study conducted in Kenya in 2008 where 320 students aged 10-12 years were randomly allocated to a school meal program, and a reduction in severe stunting prevalence of 15.6 percentage points (pp) (from 25.0% to 9.4%) was found for children receiving school meals.^9^ The study also provided information on the reduction in moderate stunting prevalence of 7.5 pp (from 28.1% to 20.6%), but we conservatively considered only the impact on severe stunting. A second study, also from Kenya, followed students at a local primary school with an average age of 5.8 years. The authors compared a control group (81 students) and a group participating in school feeding programs for 1 year (67 students); they estimated a reduction in stunting prevalence of 10 pp (from 22% to 12%) for those receiving school meals.^6^ A third study considered was a comparative cross-sectional study conducted in 2021 in Ethiopia, with a total of 936 primary school students involved. A reduction in stunting of 7.9pp (from 21.6% to 13.7%) for those receiving school meals was found ^10^. Considering two additional studies that found no effect on stunting through school meals, we assumed a reduction in stunting of 24%, taking the average of the impact retrieved from all relevant studies identified in our review.

We used a disability weight for stunting of 0.002 and assumed the disability to last from onset age to the expected end of life, based on the life expectancy at age 6 in each country. For the calculation of years of life lost, we used the generic mortality rate for children aged 5-14 years^11^ and adjusted it for the increased risk (hazard ratio of 1.47) of dying if suffering from stunting,^12^ and using life expectancy at the corresponding age of death. The formula for calculating the mortality when suffering from stunting (*S*) is reported below, where *M* is the mortality rate for children aged 5-14, *P* is the prevalence of stunting, and *R* is the hazard ratio for stunting:

$S=\frac{M}{P*\left( 1-\frac{1}{R} \right)+(\frac{1}{R})}$ .

*Diarrheal diseases*

For the impacts on diarrheal diseases, we assumed a reduction of 27% in incident cases for those provided with education on handwashing with soap before eating or food handling, after defecation, or a combination of these.^14^ This number is based on a systematic review that looked at studies from Africa and Asia conducted up to the year 2016 on under-five years old children. For YLD calculations, we used a disability weight of 0.170.^15^

### Appendix C: Effects on education and health

**Table C1**: Summary of educational outcomes by country^16^

| **Country** | **Average additional years of schooling** | **Percentage increase in enrollment** |
| --- | --- | --- |
| **Burundi** | 1.8 | 40% |
| **Ethiopia** | 2.3 | 37% |
| **Malawi** | 0.7 | 10% |
| **Mozambique** | 0.6 | 9% |
| **Namibia** | 1.5 | 31% |

**Table C2:** Summary of health-specific outcomes, by country

| **Country** | **Cases averted (per 1,000 children)** | | |
| --- | --- | --- | --- |
|  | **Anemia** | **Stunting** | **Diarrhea** |
| **Burundi** | 1,418 | 139 | 1,672 |
| **Ethiopia** | 475 | 46 | 992 |
| **Malawi** | 355 | 72 | 831 |
| **Mozambique** | 1,400 | 90 | 1,100 |
| **Namibia** | 671 | 53 | 1,272 |

**Table C3**: Summary of disability-adjusted life years (DALYs) averted and associated monetary value, by country

| **Country** | **DALYs averted (per 1,000 children)** | | **Monetary value (per 1,000 children) (USD)** | | |
| --- | --- | --- | --- | --- | --- |
|  | **Considering anemia only** | **Including stunting and diarrhea** | **GDP per capita^17^** | **Anemia only** | **All health outcomes** |
| **Burundi** | 51 | 89 | 219 | 11,189 | 19,527 |
| **Ethiopia** | 5 | 35 | 1,134 | 5,670 | 39,687 |
| **Malawi** | 4 | 57 | 523 | 2,090 | 29,788 |
| **Mozambique** | 15 | 82 | 657 | 9,852 | 53,858 |
| **Namibia** | 7 | 37 | 4,413 | 30,892 | 163,285 |

**Table C4**: Alternative benefit-cost ratio (BCR) estimates, considering the broader impact on health.

| **Country** | **Total discounted cost (USD)** | **Monetary benefits per beneficiary (USD)** | | **Benefit-Cost Ratio (BCR)** |
| --- | --- | --- | --- | --- |
|  |  | **Education** | **Health** |  |
| **Burundi** | 375 | 1,160 | 20 | 3 |
| **Ethiopia** | 273 | 2,172 | 40 | 8 |
| **Malawi** | 95 | 1,138 | 30 | 12 |
| **Mozambique** | 212 | 764 | 54 | 4 |
| **Namibia** | 158 | 4,811 | 163 | 32 |

**Table C5**: Cost-effectiveness analysis estimates

| **Country** | **DALYs averted (per beneficiary)** | | **Cost per beneficiary (USD)** | **Incremental Cost-Effectiveness Ratio (USD/QALY)** | |
| --- | --- | --- | --- | --- | --- |
|  | **Considering anemia only** | **Including stunting and diarrhea** |  | **Considering anemia only** | **Including stunting and diarrhea** |
| **Burundi** | 0.051 | 0.089 | 409 | 8,013 | 4,592 |
| **Ethiopia** | 0.005 | 0.035 | 273 | 54,661 | 7,809 |
| **Malawi** | 0.004 | 0.057 | 95 | 23,689 | 1,662 |
| **Mozambique** | 0.015 | 0.082 | 212 | 14,130 | 2,585 |
| **Namibia** | 0.007 | 0.037 | 158 | 22,513 | 4,259 |

**Webappendix references**

1.	World Food Programme. *Pilot Impact Evaluation of the Commodity Voucher Procurement Model in Burundi*. 2024. Accessed April 26, 2026. https://wfp.tind.io/record/129813/files/ELR%202936%20v.1-English.pdf

2.	World Food Programme. *Annual Country Report 2023 Ethiopia*. 2023. Accessed May 5, 2024. https://www.wfp.org/operations/annual-country-report?operation_id=ET02&year=2023#/26253

3.	World Food Programme. *Malawi Annual Country Report*. 2022.

4.	Republic of Mozambique. Programa Nacional de Alimentação Escolar - PRONAE. Preprint posted online May 14, 2013.

5.	Ministry of Education Arts and Culture. Namibia School Feeding Policy 2019. *Republic of Namibia*. Preprint posted online 2019.

11.	IHME. GBD Compare. Institute for Health Metrics and Evaluation. 2026.

17.	The World Bank. GDP per capita (current US$) - Burundi, Ethiopia, Malawi, Mozambique, Namibia. Country official statistics, National Statistical Organizations and/or Central Banks. 2026.
